# Racial Differences in Negative Symptoms of Schizophrenia: Examining the Role of Defeatist Beliefs and Discrimination

**DOI:** 10.64898/2026.04.08.26350400

**Authors:** Desmond J. Spann, Lauren M. Hall, Alexandra Moussa-Tooks, Julia M. Sheffield

## Abstract

**Background:** Negative symptoms are core features of schizophrenia that relate strongly to functional impairment, yet interventions targeting these symptoms remain largely ineffective. Emerging theoretical work highlights how environmental factors may shape and maintain negative symptoms. Although racial disparities in schizophrenia diagnosis among Black Americans are well documented and linked to racial stress and psychosis, the impact of racial stress on negative symptoms has not been examined. This study provides an initial test of a novel theory proposing that racial stress - here measured by racial discrimination - influences negative symptom severity through exacerbation of negative cognitions about the self, particularly defeatist performance beliefs (DPB).

**Study Design:** Participants diagnosed with schizophrenia-spectrum disorder (SSD) (N = 208; 80 Black, 128 White) completed the Positive and Negative Syndrome Scale (PANSS), the Defeatist Beliefs Scale, and self-report measures of subjective racial and ethnic discrimination (Racial and Ethnic Minority Scale and General Ethnic Discrimination Scale). Relationships among variables were tested using linear regression and mediation analysis.

**Study Results:** Black participants exhibited significantly greater total and experiential negative symptoms than White participants with no group difference in DPB. Racial discrimination explained 46% of the relationship between race and negative symptoms. Among Black participants, higher DPB were associated with greater negative symptom severity. Discrimination was positively related to both DPB and negative symptoms. DPB partially mediated the relationship between discrimination and negative symptoms.

**Conclusions:** Findings suggest that racial stress contributes to negative symptom severity via defeatist beliefs among Black individuals, highlighting potential targets for culturally informed interventions.

## Introduction

Black Americans are diagnosed with schizophrenia-spectrum disorders (SSD) at a rate 2.42 times higher than White Americans^1,2^. While mechanisms of systemic racism, racial stress and racial bias, have been identified as drivers of these disparities in positive symptoms, similar investigations have not been conducted within negative symptoms^3–6^. This gap is critical given that negative symptoms are the strongest predictors of long-term functional impairment (e.g., avolition; anhedonia)^7^. Furthermore, emerging evidence suggests Black individuals rate higher on negative symptoms in both self-report and clinician-report assessments^8,9^. Unraveling the “why” behind these disparities is paramount to reduce inequitable, harmful treatment and assessment through identifying mechanisms of focus for more culturally informed services.

Recent work emphasizes that rather than negative symptoms being fixed traits, these symptoms arise through ongoing interactions between individuals and their environment^10^. The bioecosystem theory of negative symptoms, based on Bronfenbrenner’s ecological framework^11^, situates individuals within interconnected environment systems, from immediate social contexts to broader cultural factors, that dynamically shape symptom expression. This work is motivated by wanting to better understand factors that could increase the efficacy of psychosocial interventions for these symptoms. Framed within this model, racism can represent a powerful environmental influence for Black individuals with the potential to exacerbate symptoms or impact interpretation of behaviors that mimic negative symptoms.

Racism is a significant environmental factor shaping behavior that Black Americans experience more than any other racial minority^12–15^. It functions across multiple interrelated levels (e.g., internalized, interpersonal, instructional and structural, and cultural) that each impose stressors that erode well-being through exposure to various direct and in-direct discriminatory experiences. The cumulative effects of these experiences, termed racial stress, tax psychological and social resources and are robustly linked to increased anxiety, depression, PTSD, and psychosis symptoms among Black Americans^16–18^. Given the widespread effects and the bioecosystem framework, it is plausible that racism contributes to the development and assessment of negative symptoms in schizophrenia-spectrum disorders.

Well-established cognitive models of negative symptoms propose that maladaptive self-concept and self-schemas, evidenced by low self-esteem and self-efficacy and increased self-defeatist beliefs, maintain symptoms by reducing motivation and reinforcing withdrawal^19–25^. Racism disrupts adaptive self-schema development and contributes to formation of beliefs that undermine self-worth, social-value, and perceived competence^26–27^. Racism, especially internalized racism, has been linked to lower self-esteem and greater psychological distress suggesting a cognitive pathway (e.g., individual factor in the bioecosystem framework) through which racial stress can exacerbate negative symptoms^28–32^. This pathway aligns with recent conceptual work proposing that racial stress may influence the severity and perception of negative symptom in Black individuals^33^.

The present study aimed to provide further evidence of disparities in negative symptom severity for Black individuals and clarify potential driving factors underlying these disparities by investigating whether maladaptive beliefs, specifically defeatist performance beliefs (DPB), mediate the relationship between racial discrimination and negative symptom severity among Black participants. We hypothesized that (1) Black Americans would have more severe negative symptoms compared to White Americans; (2) greater discrimination would be associated with higher DPB and negative symptoms; (3) discrimination would mediate the relationship between race and negative symptom severity; and (4) DPB would mediate the relationship between discrimination and negative symptoms. This work provides an initial empirical test of a recently proposed theoretical framework linking racial stress, cognitive vulnerability, and negative symptoms expression and interpretation to contribute to a more culturally informed understanding of schizophrenia-spectrum disorders^33^.

## Methods

### Participants

Adults with SSD were recruited as part of two protocols (IRB# 201882; 202462) at the Vanderbilt University Medical Center (VUMC). SSD participants were recruited through clinician-referral at Vanderbilt Psychiatric Hospital, VUMC and related clinics, direct in-house medical record review, and a community mental health center in Nashville, Tennessee. Both protocols specifically recruited SSD patients with either current or recent delusional thought content and included participants regardless of their negative symptom severity. Additional inclusion criteria were ages 18-65, lack of significant neurological disorder (e.g., epilepsy), and no history of autism spectrum disorder or traumatic brain injury. The final combined sample for this investigation included 208 participants (n = 80 Black, n = 128 White) who met DSM-5 diagnostic criteria for SSD. Table 1 displays demographic information of the sample.

**Table 1.**
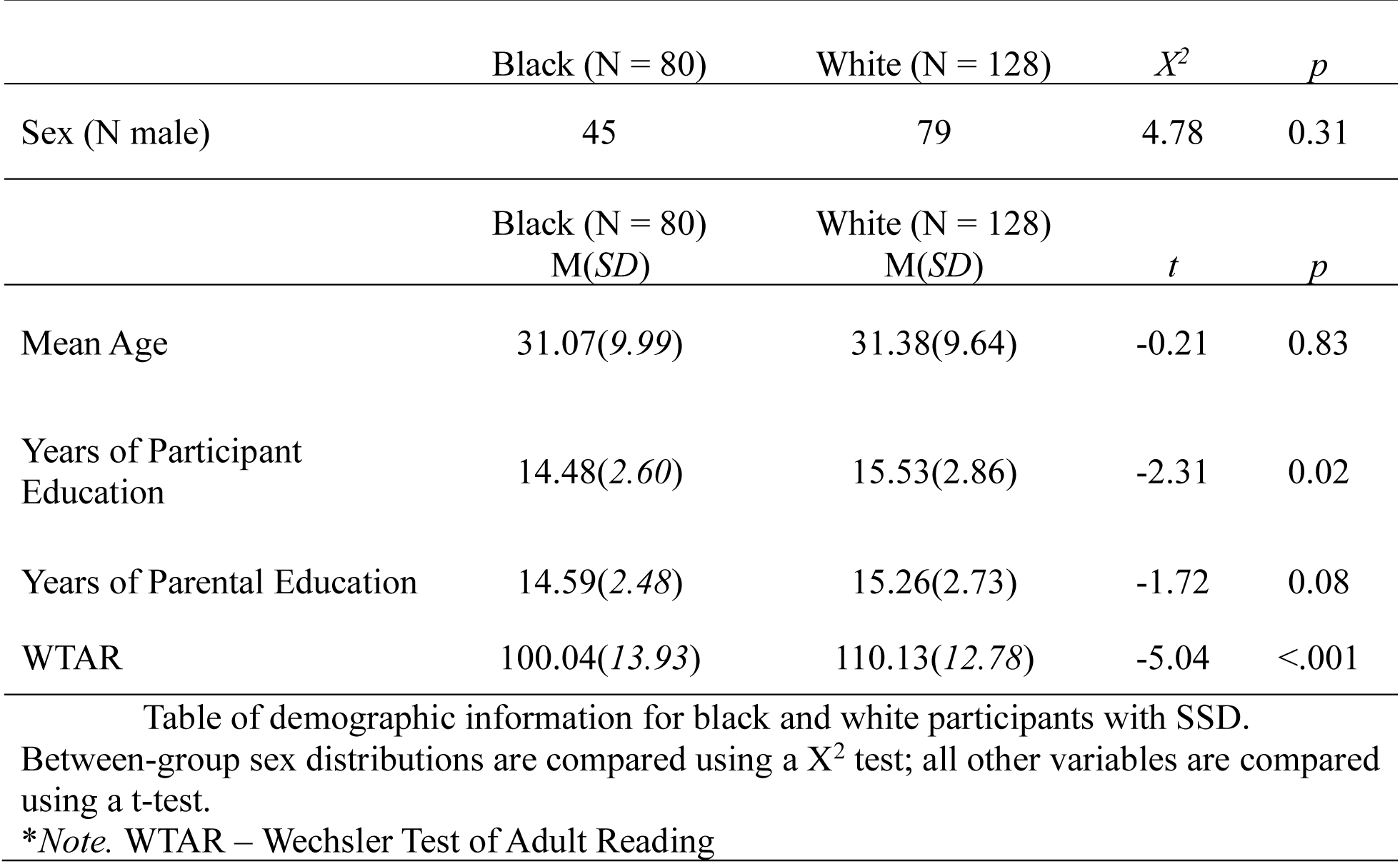
Demographics.

Table of demographic information for black and white participants with SSD. Between-group sex distributions are compared using a $X^2$ test; all other variables are compared using a t-test.
| | Black (N = 80) | White (N = 128) | $X^2$ | $p$ |
| --- | --- | --- | --- | --- |
| Sex (N male) | 45 | 79 | 4.78 | 0.31 |
| | Black (N = 80)<br>M(*SD*) | White (N = 128)<br>M(*SD*) | $t$ | $p$ |
| Mean Age | 31.07(9.99) | 31.38(9.64) | -0.21 | 0.83 |
| Years of Participant Education | 14.48(2.60) | 15.53(2.86) | -2.31 | 0.02 |
| Years of Parental Education | 14.59(2.48) | 15.26(2.73) | -1.72 | 0.08 |
| WTAR | 100.04(13.93) | 110.13(12.78) | -5.04 | <.001 |

\**Note.* WTAR – Wechsler Test of Adult Reading

### Measures

#### Negative symptoms

Negative symptoms were assessed with the Positive and Negative Symptom Scale (PANSS) which is a semi-structured clinician interview that includes 30 items assessing positive symptoms, negative symptoms, and ‘general’ symptoms, each rated on a 7-point scale (1 = absent, 7 = extreme severity)^33^. Based on previous analysis, both abstract thinking and stereotyped thinking were dropped from analysis to increase reliability of the negative symptom scale^34^. Furthermore, negative symptom domains were calculated and labeled as “experiential” (Emotional withdrawal, passive social withdrawal, active social avoidance) or “expressive” (Blunted affect, poor rapport, lack of spontaneity and flow of conversation, motor retardation). Subscale scores were created by summing the items in each domain.

#### Defeatist Beliefs

Defeatist beliefs were measured using the Defeatist Performance Beliefs scale (DPB) which assesses the participant’s beliefs and attitudes about their ability to perform goal-directed tasks (e.g., “if you cannot do something well, there is little point in doing it at all; if I fail partly, it is as bad as being a complete failure”)35. The DPB included 5 items and is rated using a 7-point Likert scale ranging from 1 (Totally Agree) to 6 (Totally Disagree) with lower scores indicating higher defeatist beliefs. The analysis included summed scores for all participants.

#### Discrimination

Experiences of discrimination were assessed with the combination of two measures. First, the General Ethnic Discrimination Scale (GED) is an 18-item measure of perceived ethnic discrimination that assesses frequency of discriminatory events over the last six months using a 6-point Likert scale ranging from 1 (Never) to 6 (Almost all the time)37. Second, the Racial and Ethnic Microaggression Scale (REMS) is a 28-item scale that assesses participants’ experiences of microaggressions across six different domains (e.g., assumed criminality, inferiority, and similarity) over the last month using a 6-point Likert scale ranging from 0 (I did not experience this event during the last month) to 5 (I experienced this event 5 or more times). The GED assesses more overt instances of perceived discrimination (i.e., How often have you been treated unfairly by your employers, bosses, and supervisors because of your race/ethnic group?) while the REMS more subtle forms of perceived discrimination (i.e., I was ignored at school or at work because of my race). Higher scores on both scales indicate more discriminatory experiences.

#### Data Analysis

All analyses were conducted in R (Version 4.5.1). Data was screened for accuracy, outliers, and normality prior to analysis. Age and sex were included as covariates in our models. To test whether there were group differences in the severity of negative symptoms and defeatist beliefs, analyses of covariance (ANCOVAs) were conducted with race (e.g., Black and White) as the between subjects’ factor and each construct as the dependent variable. To further examine whether symptom expression differed by symptom domain, a repeated-measures ANOVA was then conducted to examine within-group differences between expressive and experiential negative symptoms and whether these differences varied by race. Four White participants were noted to select the top response for every item on the REMS, despite reverse scoring, suggesting they meant to select “no” to all items. These scores were changed to “zero” after review of the data and consultation with research assistants who observed this behavior. To confirm this did not impact results, analyses were rerun with these participants excluded and no changes in the pattern of results were found.

To examine the association among negative symptoms, DPB, and discrimination, a series of linear regression analysis were conducted among Black participants. DPB and negative symptoms served as outcome variables in separate models. DPB were regressed onto negative symptoms and discrimination was regressed on to both DPB and negative symptoms.

Parallel indirect effect analyses were conducted to examine (a) whether discrimination accounted for group differences in negative symptoms, and (b) whether defeatist beliefs mediated the relationship between discrimination and negative symptoms among Black participants. Mediation was tested using nonparametric bootstrapping with 5,000 resamples (via the *mediation* package in R). Indirect, direct, and total effects were estimated. We quantified indirect effects using the Average Causal Mediation Effect (ACME) that estimates the portion of the total effect of the predictor on the outcome that operates through the mediator (i.e., the size of the mediated pathway). We also calculated the Robust Effect Size Index (RESI) for mediation effects. RESI quantifies the proportion of all explainable effects that can be attributed to the mediation pathway adjusting for model strength and suppressor effects. Higher RESI values indicate that a larger share of the association between the predictor and outcome is carried through the mediator even when total effects are small or nonsignificant.

Exploratory analyses examined whether these effects differed across negative symptoms subdomains (experiential vs expressive) and distinct types of racism (e.g., general ethnic distress versus microaggressions). Separate models were run for each subdomain and Bonferroni corrections were applied to control for multiple comparisons (⍺ = .05/3 = .017).

## Results

### Group differences in negative symptoms and defeatist beliefs

Negative symptoms were significantly greater in Black participants than White participants (F (1, 163) = 7.94, p = .005, Cohen’s f = .22). A significant main effect of symptom domain (F (1, 163) = 6.60, p = .01, Cohen’s f = .20) revealed more severe experiential than expressive symptoms across both groups (Fig 1A); however, the interaction between race and symptom domain was not significant (F (1, 163) = 2.18, p = .14). Post-hoc analysis revealed that Black participants had significantly greater experiential negative symptoms (F (1, 112) = 6.17, p = .014, Cohen’s f = .17) than White participants (Fig 1: B). Group differences for expressive symptoms approached significance (F (1, 46) = 3.43, p = .066, Cohen’s f = .13), again with Black participants having higher symptoms on average.

**Figure 1.**
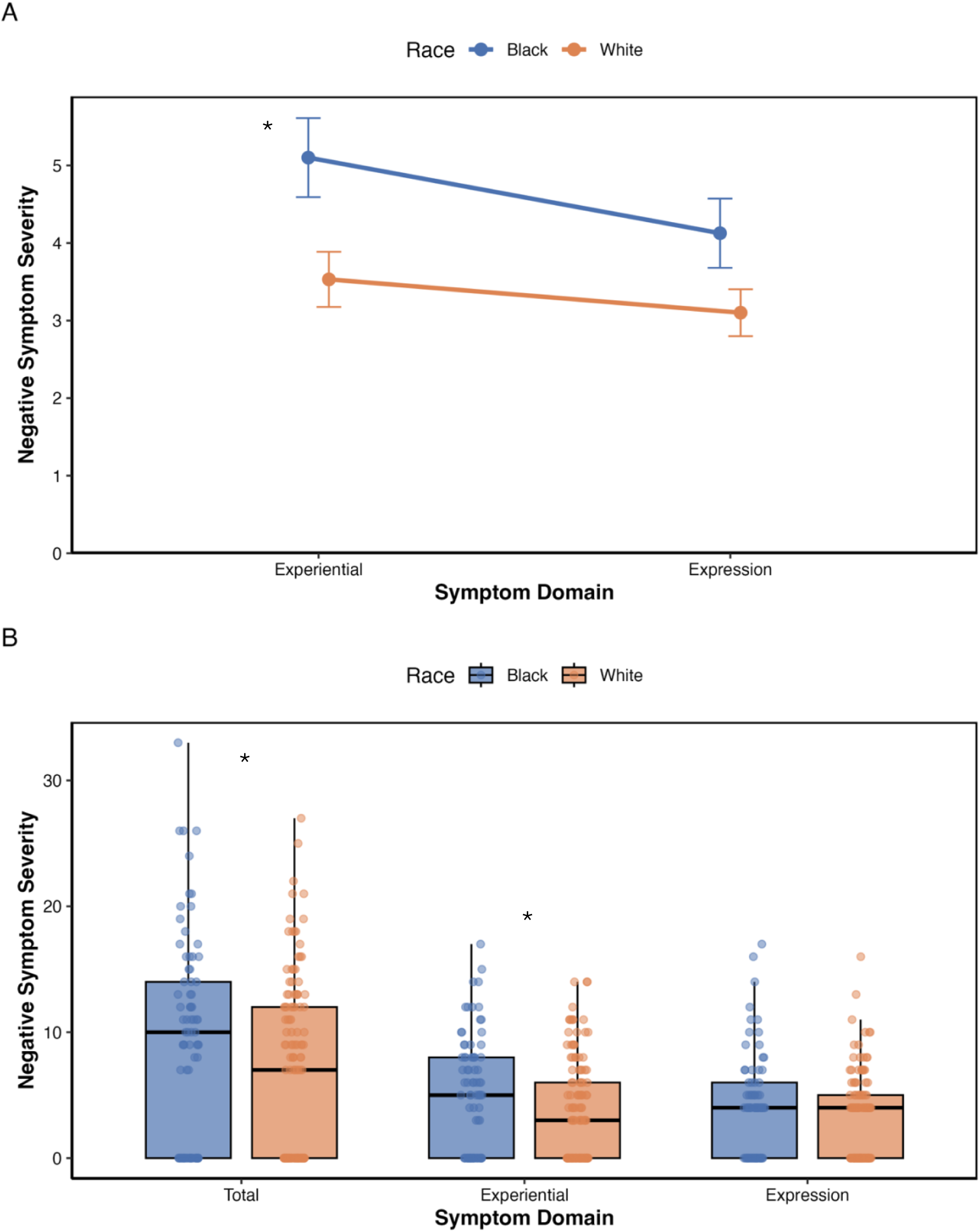
Group differences in negative symptoms; A. Repeated-measures ANOVA illustrating differences in negative symptoms domains (experiential and expression). B. Boxplots depicting the distribution of negative symptom scores across domains for Black and White participants. *Note.* * denotes significant difference.

With regards to defeatist beliefs, there were no significant group differences (F (1, 95) = 1.37, p = .244), suggesting similar self-reported defeatist beliefs in both White and Black participants.

### Relationship between defeatist beliefs, discrimination, and negative symptoms

To test our hypothesized model of the impact of racial discrimination on negative symptoms in Black participants with SSD, we conducted the remaining analyses in only those who identified as Black. Given the lack of a significant race by symptom domain interaction, we focused on total negative symptoms.

Higher defeatist beliefs were significantly associated with greater total negative symptoms (β = 0.43, p < .001, r^2^ = 0.30) suggesting that more defeatists beliefs about one’s abilities are linked to higher negative symptoms severity (Fig 2A).

**Figure 2.**
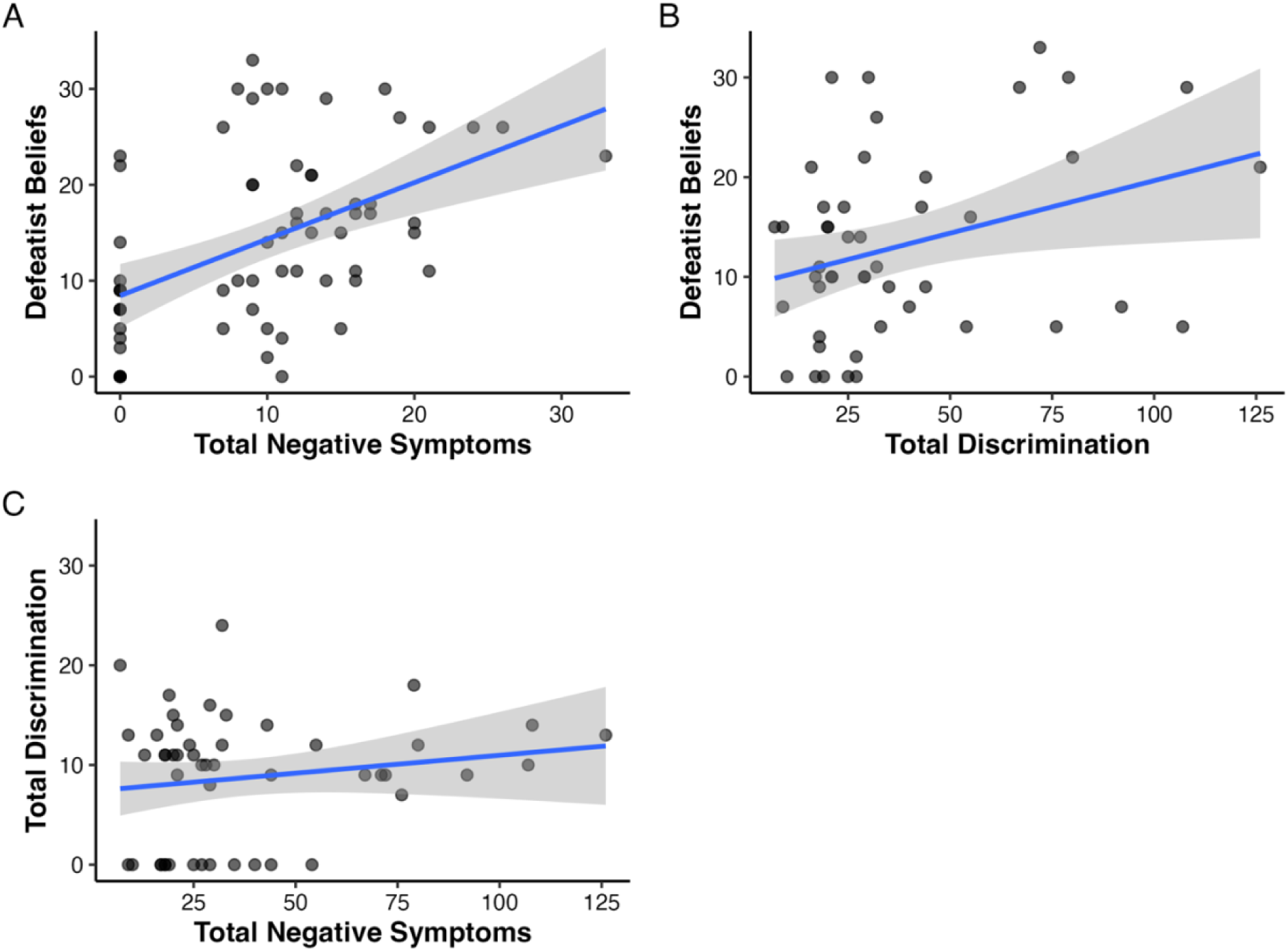
Black Participant regression models; A. Associations between defeatist beliefs and total negative symptoms, B. Associations between defeatist beliefs and total discrimination, C. Associations between total discrimination and total negative symptoms

Higher defeatist beliefs were significantly associated with greater total discrimination (β = 3.80, p < .001, r^2^ = 0.18) indicating that greater exposure to discrimination is linked to more negative or defeatist beliefs about one’s abilities among Black participants with SSD (Fig 2B). Total negative symptoms were not significantly associated with total discrimination (β = 1.20, p = 0.244) (Fig 2C).

### Mediation Analyses

Results indicated a significant indirect effect of race on negative symptoms through total discrimination (ACME estimate = −1.22, *p* = 0.01), suggesting that greater exposure to discrimination among Black participants contributed to higher negative symptoms severity (Fig 3: A). The direct effect of race on negative symptoms was nonsignificant (ADE estimate = −1.34, *p* = .32) while the total effect was significant (estimate = −2.46, *p* = 0.04). Approximately 46% of the total effect of race on negative symptoms was mediated by discrimination (*p* = 0.048) which indicates that nearly half of the racial differences in symptom severity were explained by differential exposure to discrimination. RESI further showed that 42% of the explainable variance in the racial group and symptom severity relationship operated through the discrimination pathway.

**Figure 3.**
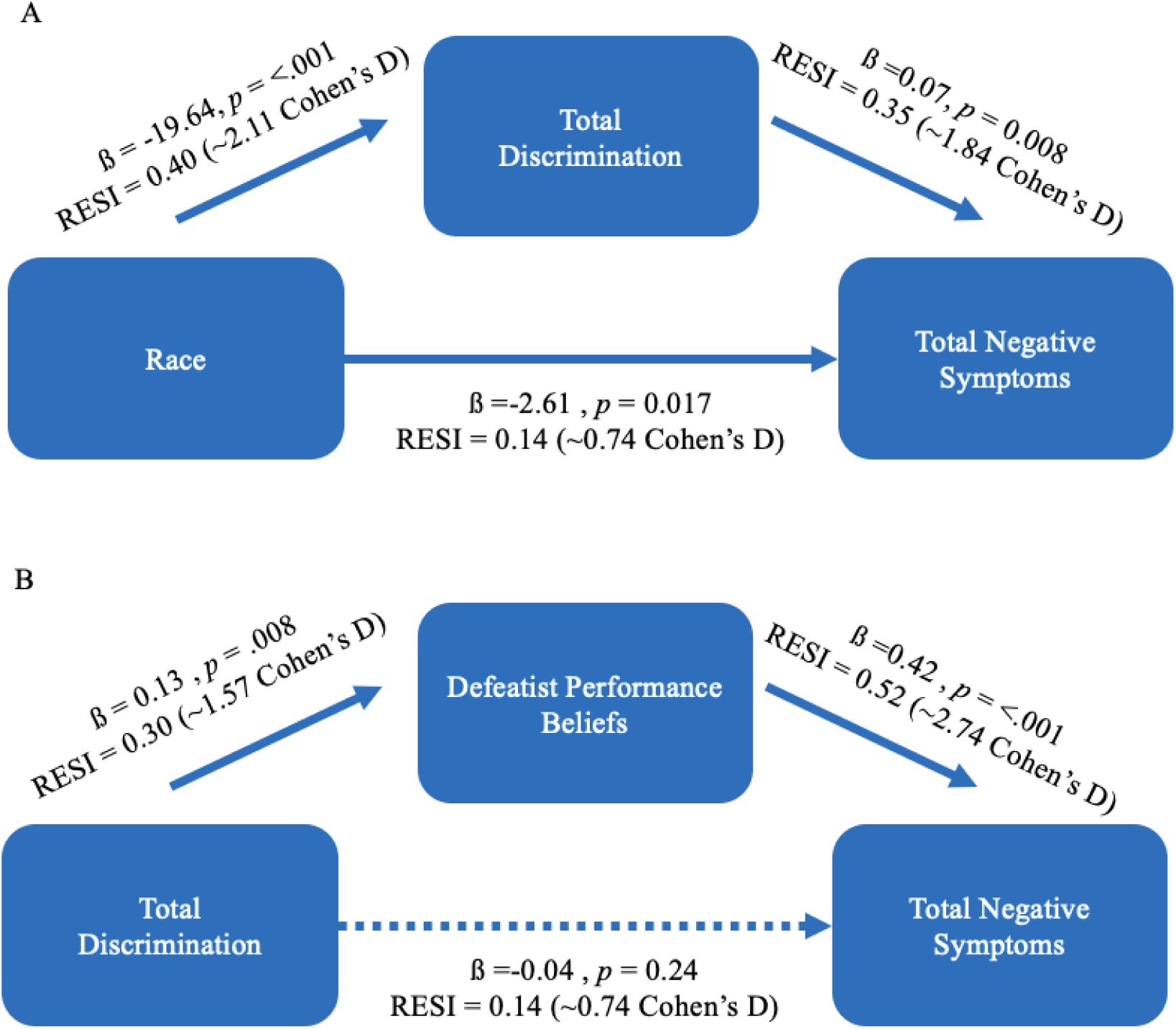
Mediation Analyses; A. Total discrimination mediating race and total negative symptoms, B. Defeatist performance beliefs mediating total discrimination and total negative symptoms. Note: Beta values are provided as well as effect size estimates using the robust effect size index (RESI) and a Cohen’s d equivalent. Solid lines indicate significant paths and dotted lines indicated non-significant paths.

Although the primary path between negative symptoms and discrimination did not reach statistical significance, we still aimed to test our *a priori* model to examine the interplay between discrimination, defeatist beliefs, and negative symptoms in our Black participants. We observed a significant indirect effect of defeatist beliefs on the relationship between total negative symptoms and self-reported discrimination (ACME estimate = 0.04, p = 0.03), indicating that defeatist beliefs explained a significant proportion of variance in the relationship between subjective discrimination and negative symptoms severity in Black participants with SSD (Fig 3: B). RESI analyses showed that defeatist beliefs accounted for 23% of the variance in this association.

### Exploratory Analyses

Exploratory associations among specific dimensions of negative symptoms (expressive and experiential) and the different forms of discrimination (general ethnic discrimination and microaggressions) were analyzed to contextualize primary findings. Across the models, the overall patterns were consistent with our main analysis. Discrimination related to greater negative symptom severity with evidence of defeatist beliefs as a mediator. Although many effects were modest or approached significance after correction for multiple comparisons, the direction and pattern of the relationships were similar across symptom domains and discrimination subtypes. Detailed results are provided in the supplement.

## Discussion

The present study sought to test the presence of racial disparities in negative symptom severity among individuals with SSD and clarify mechanisms contributing to these disparities. Black participants exhibited higher levels of total and experiential negative symptoms compared to White participants, and racial discrimination had a significant indirect effect on this relationship, accounting for nearly half of the variance in differing symptom severity between racial groups. These findings suggest the possible role of racial stress in the phenotype of negative symptoms in SSD. To further understand how this occurs, we hypothesized that discrimination may exacerbate defeatist performance beliefs– a core cognitive mechanism of negative symptoms. We found that, among Black participants, greater perceived discrimination was significantly associated with more defeatist performance beliefs, and that defeatist beliefs mediated the relationship between racial discrimination and negative symptoms. These results support cognitive models of negative symptoms while extending them within a multicultural framework, by identifying perceived discrimination as a meaningful link between race and symptom severity. Specifically, maladaptive self-schemas and defeatist beliefs may develop as cognitive responses to sustained racial stress serving as a key mechanism translating environmental stress into clinical expression.

The finding that racial discrimination explains 46% of the variance between race and negative symptom severity highlights racism as a critical social determinant for symptom expression and clinical presentation. The bioecosystem theory posits that negative symptoms can be better conceptualized through dynamic environmental interactions rather than classifying them as a fixed trait^10^. Results suggest that within the bioecosystem framework, racial discrimination represents a unique, chronic environmental stressor for Black individuals that operates across the ecological systems (e.g., internalized, interpersonal, institutional and structural, and cultural) to shape behavioral and psychological functioning. Theoretically, these dimensions of racism exist across the environmental systems the individual is embedded in and create racial stress through dynamic interactions that can negatively impact cognitive processes found within the individual. That is, this racial stress becomes internalized and influences negatively biased self-schemas^16,26^. This may be especially true for experiential symptoms (e.g. avolition, anhedonia) where these biased cognitive processes reinforce reductions in goal-oriented behavior and social withdrawal. Our findings suggest that these cognitive vulnerabilities may be, in part, socially constructed through the lived experiences of racism that systematically undermines self-worth and perceived competence among Black individuals.

Critically, negative-biased self-schemas, most notably defeatist performance beliefs, are central to current conceptualizations of negative symptoms in schizophrenia-spectrum disorders. Cognitive models of negative symptoms posit that symptoms are rooted in maladaptive appraisals of both the self and the external world where these distorted beliefs contribute to enduring negative perceptions of the self, reduced motivation, and disengagement from goal-directed and social activity^19–20^. Therefore, the finding that DPB was related to racial discrimination and significantly mediated the relationship between discrimination and negative symptoms (e.g., indirect pathway uniquely explains 23% of the variance) among Black SSD participants offer foundational support for the proposed theoretical pathway. Specifically, that racial discrimination exacerbates negative symptoms largely through its impact on maladaptive self-referential cognitions. This result reinforces the utility of the bioecosystem model by demonstrating the importance of embedding existing cognitive models of negative symptoms^20–21^ within environmental and cultural contexts. Cognitive vulnerabilities that contribute to negative symptom severity (e.g., low self-efficacy and defeatist beliefs) can be shaped and amplified by sociocultural experiences of racism rather than solely from individual level traits. Chronic exposure to discrimination can foster maladaptive self-schemas that erode perceived competence and reinforce defeatist beliefs that contribute to diminished motivation, anhedonia, and social withdrawal^28–32^. Within this context, DPB may reflect not only cognitive distortions but also internalized consequences of navigating persistent racial stress found across the multiple ecological systems. Clinically, these findings emphasize the value of not only targeting DPB within treatment, but also the importance of utilizing culturally informed interventions (e.g., racial healing framework) that meaningfully address the sociocultural origins of distress. Interventions that both modify maladaptive beliefs and validate the lived experiences of racism^39^ may offer not only short-term symptom reduction but also support the development of culturally meaningful coping strategies for managing chronic racial stress.

Importantly, we observed a nonsignificant direct effect of discrimination on negative symptoms. While initially surprising, this result emphasizes the complexity of racism’s impact on mental health and highlights the need to consider racial identity when examining these relationships. Black individuals do not experience or internalize racism uniformly and evidence points to the psychological effects of racism being shaped in part by racial identity, the extent to which race is central to an individual’s self-concept, and racial socialization, the process through which individuals learn culturally relevant values, beliefs, and coping strategies associated with their racial groups^40^. Key racial identity dimensions, such as racial centrality (the extent to which racial group membership is emphasized as part of overall self-concept), private regard (viewing one’s racial group positively), and public regard (perceptions of how others view one’s racial group) moderate the effects of discrimination on mental health outcomes^41^. Individuals high in centrality and positive private regard are more likely to attribute discrimination to structural factors rather than personal inadequacy protecting self-schemas from internalization of racial stress^42^. In contrast, low centrality or negative private regard can weaken this protective process making the individual more vulnerable to adopting maladaptive self-beliefs in response to racist encounters. Understanding these identity-based pathways is essential for clarifying when and for whom racial discrimination contributes to cognitive vulnerabilities such as defeatist beliefs, and ultimately, to negative symptom severity. Future work integrating racial identity processes will be critical for capturing the nuanced and individualized ways racism shapes clinical outcomes associated to negative symptoms.

There are several limitations of these findings to consider. First, the measures of perceived discrimination were all self-report measures which then depends on the participants ability to accurately assess their own experiences. Measures were also limited in the scope of discrimination and may not have been able to accurately represent all aspects of discrimination experienced by the participants. Second, there was no direct measure of racial stress. The analysis used participants reported number of racist encounters from both the GED and REMS to better encompass the broad nature of discriminatory encounters, but did not include the level of distress experienced by these events. Further investigations should examine stress associated to discriminatory events to provide more information about stress and development of maladaptive thoughts. Third, only White and Black individuals were examined within this study which limits the inferences that can be made about out racial groups. Future studies should expand the number of racial groups included to illustrate potential differential impact of racism on negative symptoms. Finally, racial discrimination does not only shape internal cognitions such as defeatist beliefs, but it can also evoke adaptive, self-protective behavioral responses that resemble negative symptoms (e.g., guardedness, reduced social engagement, or strategic withdrawal) but emerge from navigating racially threatening or invalidating environments43-45. This is an important distinction for future research on assessment bias of negative symptoms, that the current study was not set up to test.

As highlighted, these results have important implications for theory, assessment, and intervention. Theoretically, they provide empirical evidence for a novel theory positing that racism can impact negative symptoms through maladaptive self-referential cognitions. Future efforts should begin to include key constructs of racial identity to better understand the complexity of the relationships involved. They also extend the utility of the bioecosystem by empirically demonstrating that racial discrimination, a salient and proximal environmental factor, acts as a mechanism influencing negative symptom presentation. Clinically, the results emphasize the potential benefit of interventions targeting maladaptive cognitions (e.g., low self-efficacy, defeatist beliefs) especially when adapted to incorporate the role of racial stress for Black Americans with SSD.

## Data Availability

All data produced in the present work are contained in the manuscript

## Supplement

### Exploratory Analyses

Although our primary model focused on overall negative symptom severity and subjective discrimination, we also explored relationships between facets of these experiences – expressive/experiential negative symptoms and general ethnic discrimination/microaggressions.

### Experiential

Experiential symptom severity was significantly associated with DPB (β = 0.25, p < .001, R² = 0.36) while associations between total discrimination (β = 1.24, p = .03, R² = .10) and general ethnic discrimination (β = 0.09, p = .03, R² = .10) reached trend-level after multiple comparisons correction.

Discrimination significantly mediated the relationship between race and experiential negative symptoms (ACME = −0.85, p = .004) and accounted for 53% of the total effect. The direct effect was nonsignificant (ADE = −0.76, p = .299) indicating that racial differences in experiential symptoms were largely explained by discrimination.

Mediation analysis with total discrimination revealed a pattern of trend-level effects including the indirect effect (ACME = 0.03, p = .03), the total effect (estimate = 0.04, p = .03), and proportion mediated (estimate = 0.55, p = .04). This suggests that over half of the association between discrimination and experiential symptoms operated through defeatist beliefs.

### Expression

Expression symptom severity was significantly associated with DPB (β = 0.18, p < .001, R² = 0.19). The mediation analysis for total discrimination was also trend-level after multiple comparisons correction (ACME = 0.01, p = .04).

### General Ethnic Distress

GED was significantly associated with DPB (β = 0.24, p = .011, R² = 0.16) and when examined the a mediation analysis for total negative symptoms, the indirect effect (ACME; estimate = 0.07, p = .06), proportion mediated (estimate = 0.74, p = .07) and direct effect (estimate = 0.09, p = .02) approached significance when correcting for multiple comparisons.

### Microaggressions

When examining microaggressions as assessed by the REMS, indirect effects via defeatist beliefs approached significance across total (ACME; estimate = 0.05, p = .04) and expressive negative symptom domains (ACME = 0.02, p = .05) following correction for multiple comparisons. For experiential symptoms, the indirect effect (ACME = 0.03, p = .05) and direct effect (estimate = 0.05, p = .08) both trended toward significance.

Taken together, these exploratory results suggest that defeatist beliefs significantly mediate the relationship across discrimination and negative symptoms subtypes. Though some effects only approached significance after corrections, the overall pattern supports the proposed model and may reflect limited statistical power rather than absence of a true effect.

